# Indomethacin Use for Mild & Moderate hospitalised Covid-19 patients: An open label randomized clinical trial

**DOI:** 10.1101/2021.07.24.21261007

**Authors:** Rajan Ravichandran, Surapaneni Krishna Mohan, Suresh Kumar Sukumaran, Devakumar Kamaraj, Sumetha Suga Daivasuga, Samson Oliver Abraham Samuel Ravi, Sivakumar Vijayaraghavalu, Ramarathnam Krishna Kumar

## Abstract

**Introduction:** Indomethacin, a non-steroidal anti-inflammatory drug (NSAID), has been presented as a broad-spectrum antiviral agent. This randomised clinical trial in a hospital setting evaluated the efficacy and safety of this drug in RT-PCR-positive coronavirus disease 2019 (COVID-19) patients.

**Materials & Methods:** A total of 210 RT-PCR-positive COVID-19 patients, who provided consent were allotted, to control or case arm, based on block randomisation. The control arm received standard of care comprising paracetamol, ivermectin, and other adjuvant therapies. The patients in the case arm received indomethacin instead of paracetamol, with other medications retained. The primary endpoint was the development of hypoxia/desaturation with SpO_2_ ≤ 93, while time to become afebrile and time for cough and myalgia resolution were the secondary endpoints.

**Results:** The results of 210 patients were available, with 103 and 107 patients in the indomethacin and paracetamol arms, respectively. We monitored patient profiles along with everyday clinical parameters. Blood chemistry at the time of admission and discharge was assessed.

As no one in either of the arms required high-flow oxygen, desaturation with SpO_2_ level of 93 and below was an important goal. In the indomethacin group, none of the 103 patients developed desaturation. On the other hand, 20 of the 107 patients in the paracetamol arm developed desaturation. Patients who received indomethacin also experienced more rapid symptomatic relief than those in the paracetamol arm, with most symptoms disappearing in half the time. 56 patients out of 107 in the paracetamol arm had fever on the seventh day, while no patient in the indomethacin group had fever. Neither arm reported any adverse event. The fourteenth-day follow-up revealed that the paracetamol arm patients had faced several discomforts, including myalgia, joint pain, and tiredness; indomethacin arm patients mostly complained only of tiredness.

**Conclusion:** Indomethacin is a safe and effective drug for treating patients with mild and moderate covid-19.

## Introduction

SARS-Cov-2, a member of the coronavirus family, has been ravaging the world for the past 18 months. Although an effective treatment has eluded the medical community, there have been several registered trials on finding new or repurposed drugs. Several studies have discussed the mechanism of the virus-host interaction and possible treatments [1] but a safe and effective treatment for the disease is yet to emerge. Drug repurposing seems to be an immediate solution and various drugs have been suggested for COVID-19 treatment [2, 3].

The drugs required to combat the pathogen may fall into one or more of the following categories: antivirals, anti-inflammatory agents, and supporting therapies [4, 5]. According to V’Kovski, [1] the antiviral action can be based on the stages of viral-host interactions. These include attachment and virus neutralisation, host protease inhibitors that stop the entry of the virus, viral protease inhibitors, viral RdRp inhibitors, and viral maturation inhibitors. Frediansyah et al. [6] enumerated the possible antiviral solutions at various stages of interactions. The role of cathepsin L in cleavage of the S protein complex and subsequent release of virus genome is well documented. Inhibiting cathepsin L inhibits the entry of SARS-Cov-2 by 76% [7].

Pro-inflammatory cytokine production is natural during immune response. The elimination of virus-infected cells is an important step in disease control. If this step, which naturally follows virus entry and replication, is defective or prolonged, it can result in a cytokine storm,[8] an uncontrolled release of pro-inflammatory cytokines. Several interleukins are involved in a cytokine storm, the foremost being interleukin 6 (IL-6), IL-1,[8] and IL -17 [9]. IL-17 also seems to have a role as the interaction partner of SARS-Cov-2. Anti-inflammatory drugs targeting the production of these interleukins are important for COVID-19 treatment.

### Indomethacin as a Drug for SARS-Cov-2

Amici et al. [10] were the first to identify the antiviral activity of indomethacin. They recorded the antiviral activities of indomethacin against SARS-Cov-1 in vitro. Xu et al. [11] presented evidence of its antiviral activity against SARS-Cov-2. Their investigations covered the antiviral effect of indomethacin in vitro, cellulo, corona-infected canine models. They also stated that indomethacin does not reduce infectivity, binding, or entry into target cells. This conclusion is based on the results of Amici et al. [10] for SARS-Cov-1, although computer models have indicated otherwise [12]. Downregulation of ACE2 and TMPRSS2 is important to reduce infectivity. Using an open-source code, Gene2Drug, Napolitano et al.,[12] showed in a computer model that indomethacin downregulates ACE2 by suppressing the genes in the ACE2 pathway. Raghav et al. [13] depicted the role of indomethacin in inhibiting cathepsin L activity required for fusion. Interestingly, no other non-steroidal anti-inflammatory drugs [13] projected this quality.

Non-Structural Protein7 (Nsp7), along with Nsp12, is important for RNA synthesis, as highlighted by Frediansyah et al. [6], Gordan et al. [9] recognised that prostaglandin E synthase 2 (PGES-2) is an “interactor” with Nsp7, and indomethacin inhibits PGES-2. Hence, indomethacin is an important candidate for blocking RNA synthesis. Amici et al. [10] reported of this block of RNA synthesis. Amici et.al [14] also demonstrated that protein kinase R (PKR) activation by indomethacin results in inhibition of virus protein translation. This follows the work of Brunelli et al. [15], where the role of indomethacin in activating PKR directly has been demonstrated.

Several publications [16, 17] have highlighted the importance of preventing inflammation in Covid-19 patients. Indomethacin downregulates IL-6 by inhibiting the synthesis of PGES-2 [18]. Indomethacin has been successful in preventing cytokine reactions in kidney transplant patients receiving OKT3 therapy [19, 20].

Rajan et al. [21] conducted one of the first indomethacin trials. Using the data from an open-label single-arm data for indomethacin, they showed the effect of indomethacin as a treatment option by matching propensity score with retrospectively collected data on paracetamol. Gordon et al. [9] showed by retrospective data analysis that indomethacin markedly reduces the need for hospitalisation. Two studies [22, 23] have shown the effectiveness of indomethacin in treating a small number of SARS-Cov-2 patients with severe comorbidities. However, these were small case series, and a larger controlled trial is required to validate these findings.

The primary objective in this study is to determine the percentage of desaturating patients. As a quantitative criteria SpO_2_ ≤ 93 has been used as a measure. The secondary outcome was symptomatic relief. Time to become afebrile, relief from cough and myalgia are the major symptoms for the secondary outcome. The safety profile of indomethacin was also monitored as a secondary outcome.

## Methods

### Study Design

This open label randomized clinical trial consisted of two parallel groups; the control group with paracetamol and standard of care (SOC) and the indomethacin group with SOC. We recruited RT-PCR positive covid-19 patients using four and six block randomisation parallel group protocol [24]. The protocol was computer generated and all care were taken to conceal the allocation of patients from the treating physicians. They were not involved either in the randomisation or the allocation. The randomization was handled by a third person, not involved either in the treatment or in data collection. To understand the impact of the sample size, we assumed the response rate for paracetamol and indomethacin to be 0.82 and 0.95, respectively. The sample size was calculated using R with an alpha value of 0.05. The marginal power was of 0.8 [25]. The sample size worked out to be 95 in each group. The study was conducted at Panimalar Medical College, Chennai, India, in a designated Covid ward. Fig. 1 describes the study design.

**Fig. 1.**
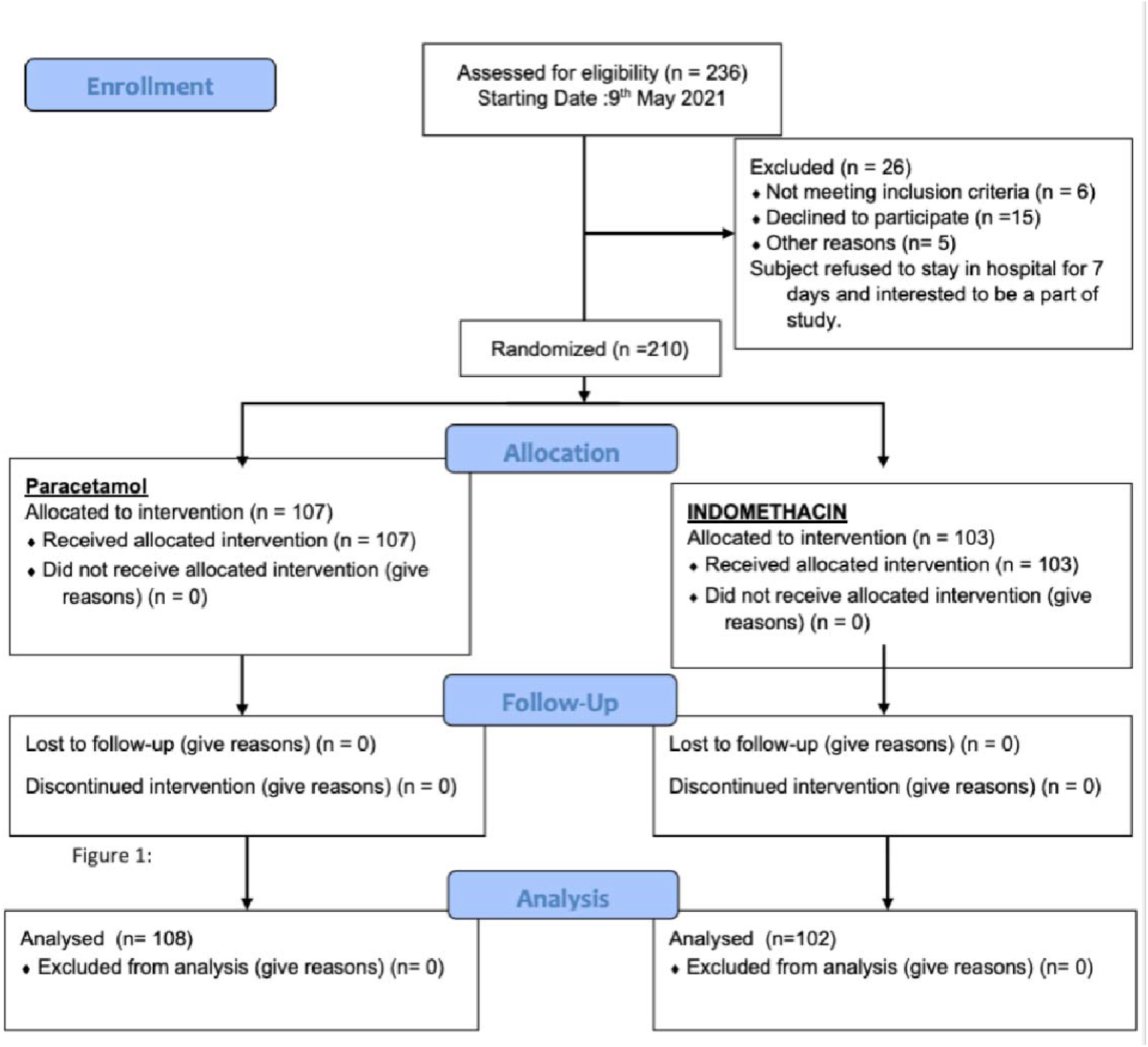
Flow Diagram

### Study Cohort

The patients recruited were RT-PCR positive. The clearly defined inclusion and exclusion criteria are given below.

Inclusion

- Age between 20 and 90 years
- RT-PCR positive
- Hospitalised patients
- The case criteria for the study:
- Oxygen saturation – 94 or more

Exclusion

- Hypersensitivity/Allergy to drugs
- Gastritis
- Recent heart attack
- Severe asthma
- Acute kidney injury
- Patients on immunosuppressants
- Pregnant & lactating mothers
- Indomethacin allergy

The trial was approved by the Panimalar Medical College Hospital & Research Institute - Institutional Human Ethics Committee with a CDSCO Registration Number: ECR/1399/INST/2020. The approval Number for the trial is PMCH&RI/IHEC/2021/051. The trial was also registered with Clinical Trial Registry of India (CTRI/2021/05/033544) of the ICMR.. The patients were apprised of the trial and the background before obtaining their informed consent which were made available in English and the vernacular language.

### Treatment

Indomethacin replaced paracetamol and was given along with the hospital standard care, which included doxycycline and ivermectin. The standard of care was decided by the government regulated protocol, followed in many hospitals throughout India. However, studies have shown that ivermectin may not be effective in treating covid-19 patients [26]. We also added a proton pump inhibitor along with indomethacin. The drugs and their dosage are listed in Table 1.

**Table 1.**
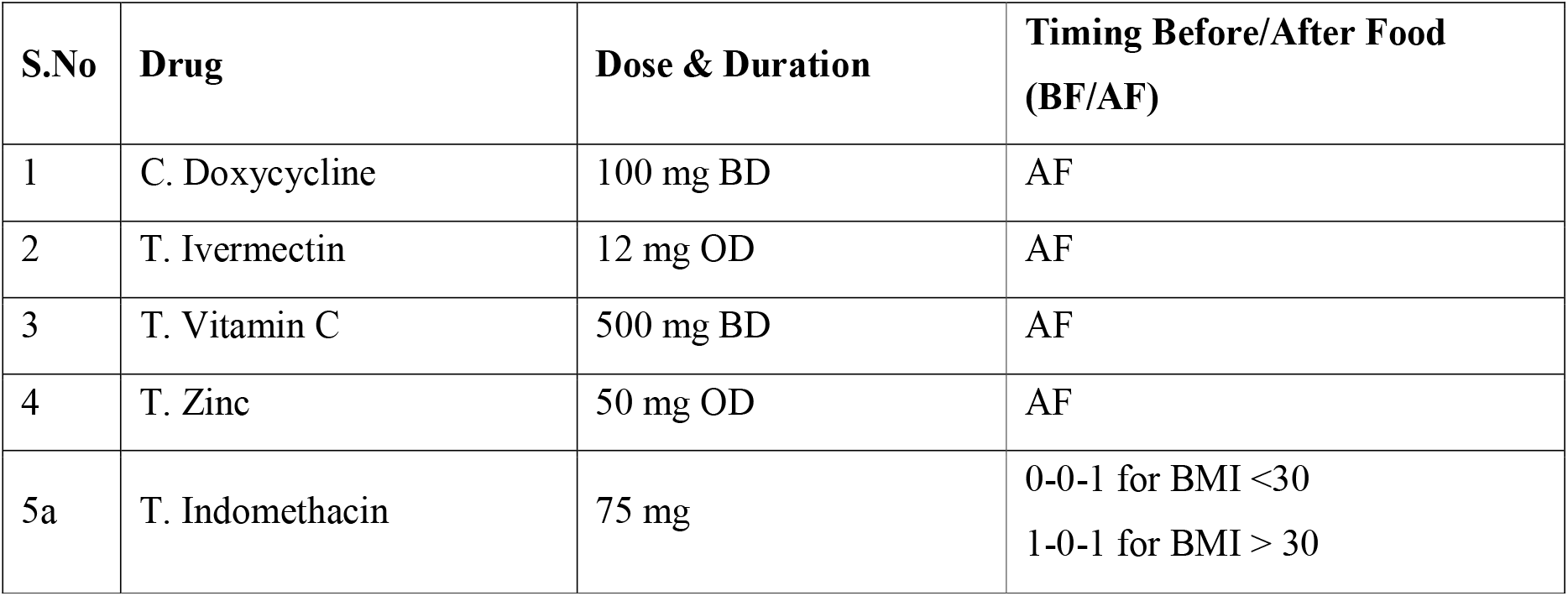
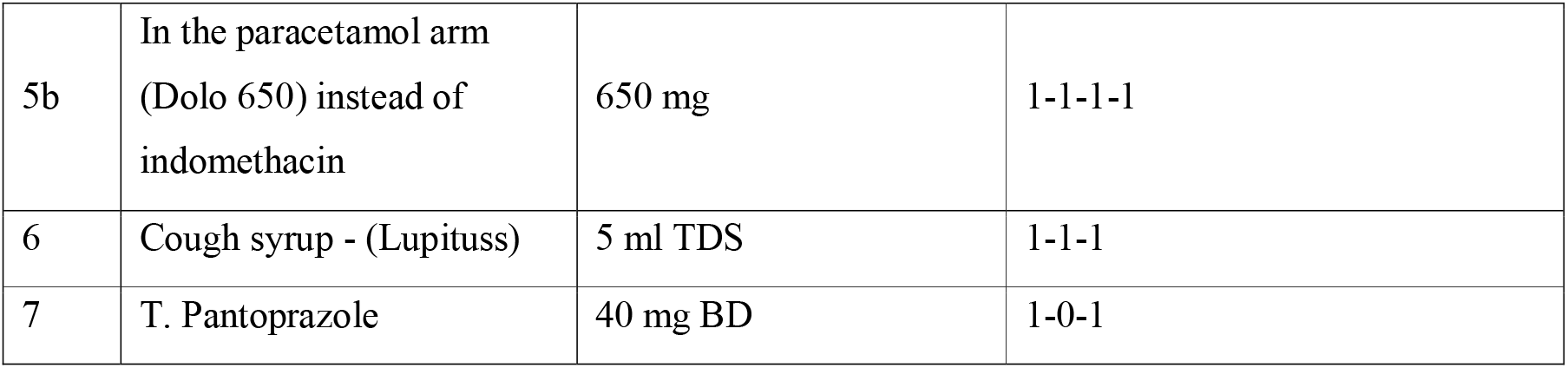
Drug chart

The treatment regimen was for five days.

### Measurements

The following investigations were conducted on admission: CT scan of the lungs, liver function test (LFT), kidney function test (KFT), C-reactive protein (CRP), and D-dimer as well regular blood tests such as complete blood count. We repeated blood chemistry on discharge and monitored the well-being of the patients after discharge for 14 days through telephonic communication. We also monitored the patients for clinical symptoms such as oxygen saturation, fever, cough, and myalgia for seven days at the hospital. Patients were deemed symptomatically recovered if the temperature dropped below 99^0^ F for two days and cough reduced to a score 1 on a one-to-ten scale (1: no cough; 2-3: cough sometimes; 4-6: coughing with the ability to do things; 7-8 persistent cough; 9-10: great deal of discomfort). RT-PCR was mandatory on admission and for 122 patients, RT-PCR was repeated on the seventh day, before discharge.

### Statistical Analysis

We used standard statistical parameters to analyze the recruited patients. The patients in the two arms were also compared based on mean/median, interquartile range (IQR) and the Wilcoxon test to estimate the p-values. Wherever appropriate 95 % confidence interval (CI) was calculated. The test of significance for p-value was based on a p-value of 0.01. We followed the dictum “once randomized always analyzed”. The primary end point was examined using chi-squared test and the secondary end points were also studied based on mean, IQR and p-values calculated using Wilcoxon test.

We used linear regression and non-linear regression to analyze the reduction in C-Reactive protein. To comprehend the time for symptomatic relief clearly, we also carried out Kaplan-Meir survival analysis and used Cox regression to understand the effect of covariates. These two results are presented in Supplementary Section.

## Results

Our goal in this study was to recruit 300 patients. The results presented here are for the first 210 patients and is an interim report. As Randomization resulted in 107 patients in the paracetamol group and 103 patients in the indomethacin group.

### Patient Characteristics and Disposition

Patient profiles are shown in Fig. 2 and Table 2. The age profile and gender-wise enumeration match closely in both groups. The temperature on admission had a marginal bias, being higher for patients in the indomethacin group. In addition, more patients in the indomethacin group had severe cough (above Scale 7). The co-morbidity distribution was similar. No patient was lost in the follow up.

**Fig. 2.**
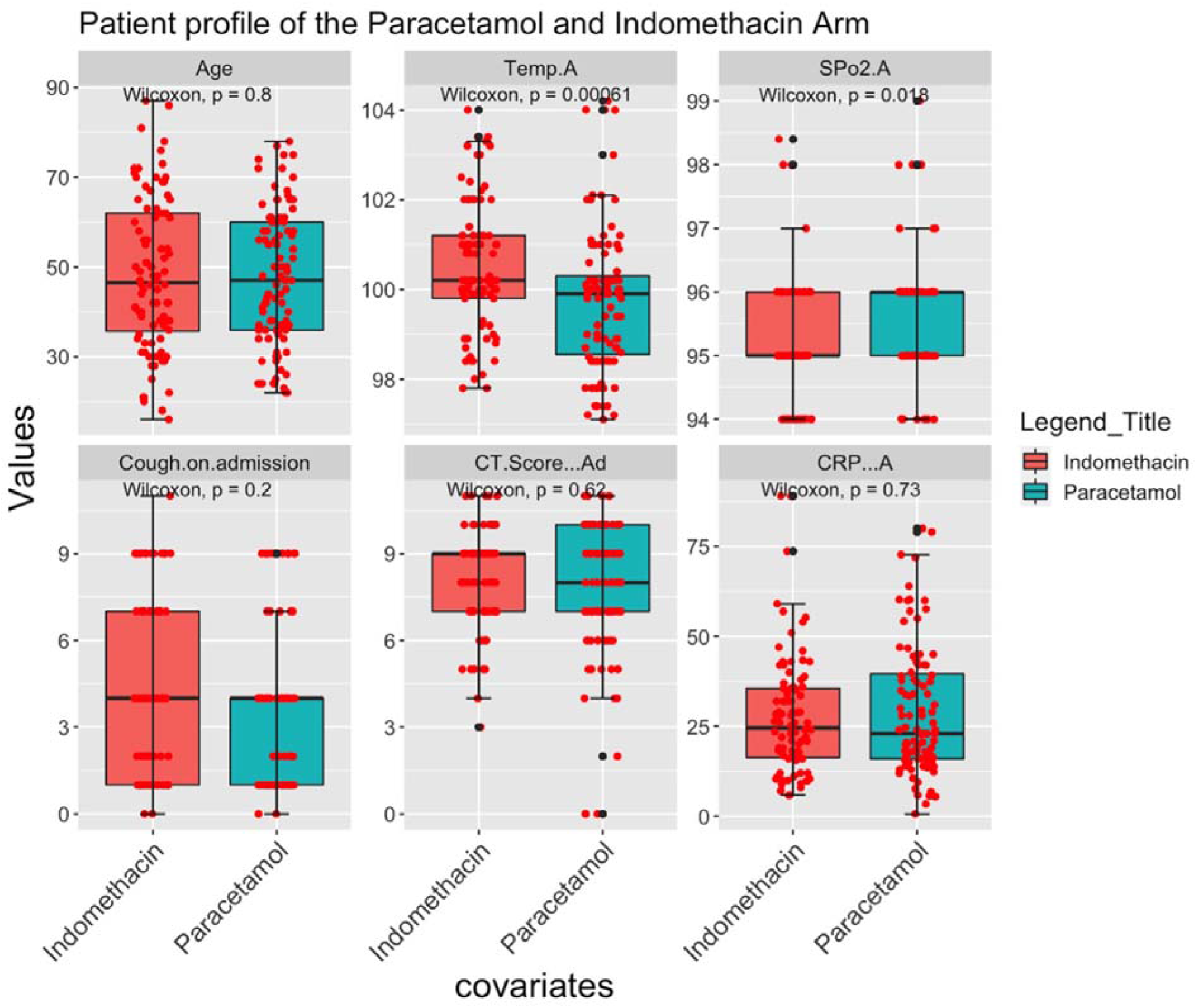
Patient Profile on admission

**Table 2.**
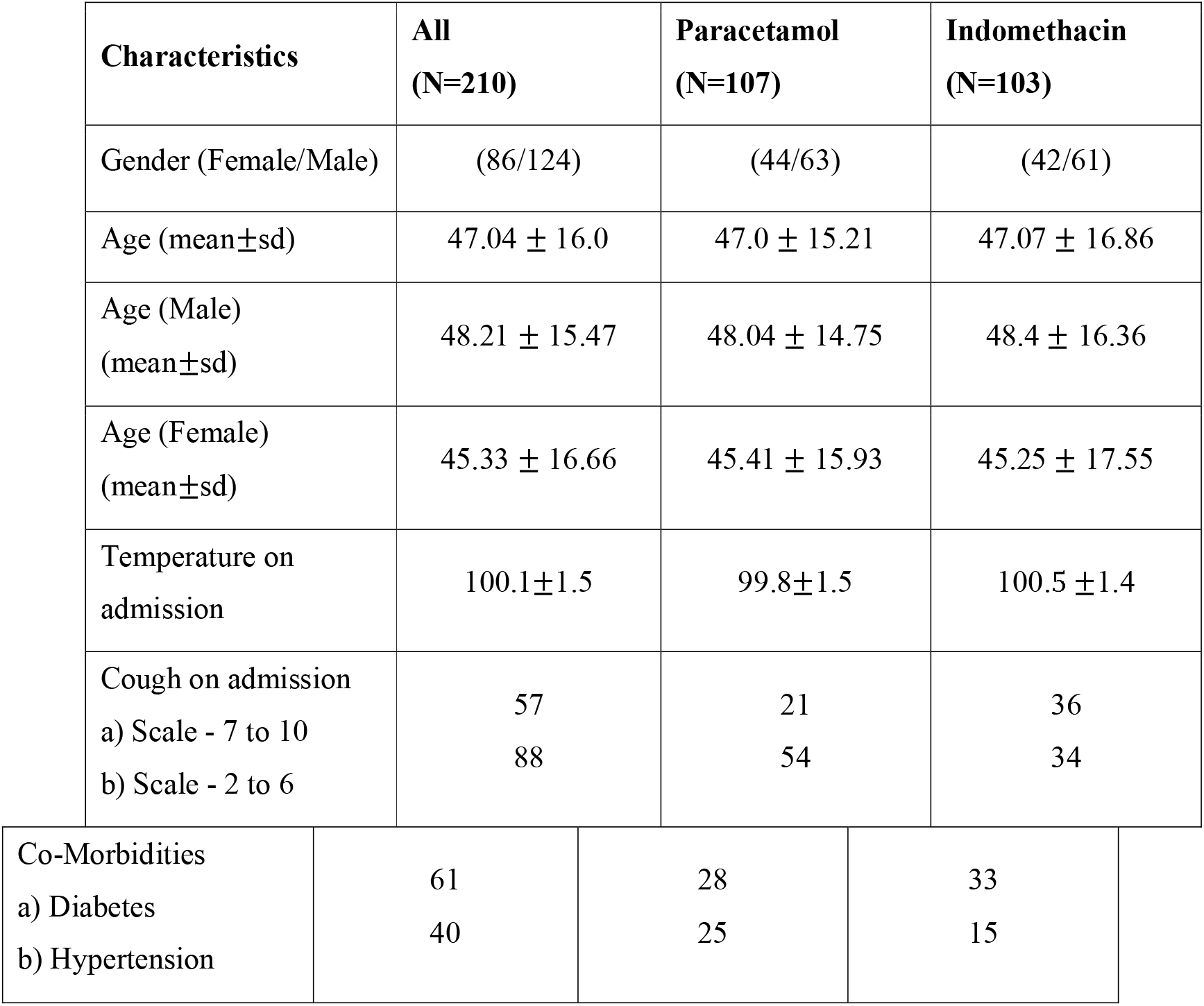
The profile of the recruited patients

### Efficacy Analysis

Symptomatic relief is very important for the psychological and physiological well-being of patients. We monitored the number of days to become afebrile, days for reduction of cough, and relief from myalgia. The results are presented in Fig. 3a for afebrile, Fig. 3b for number of days for cough reduction, Fig. 3c for cough reduction with a higher score on admission, Fig. 3d with lower cough score on admission and Fig. 3e for myalgia reduction. Median (dark line) and interquartile ranges are shown as boxes. The symptomatic recovery from fever and cough in terms of median values are given in Table 3. Two points are significant from the table and figure. Symptomatic relief with indomethacin takes only half the time compared to paracetamol. Additionally, the IQR, a measure of statistical dispersion, is very small with indomethacin compared to that of paracetamol. This is significant because the action of indomethacin is almost independent of the condition of patient on admission. The p-value in the figure indicates statistical significance.

**Fig. 3a.**
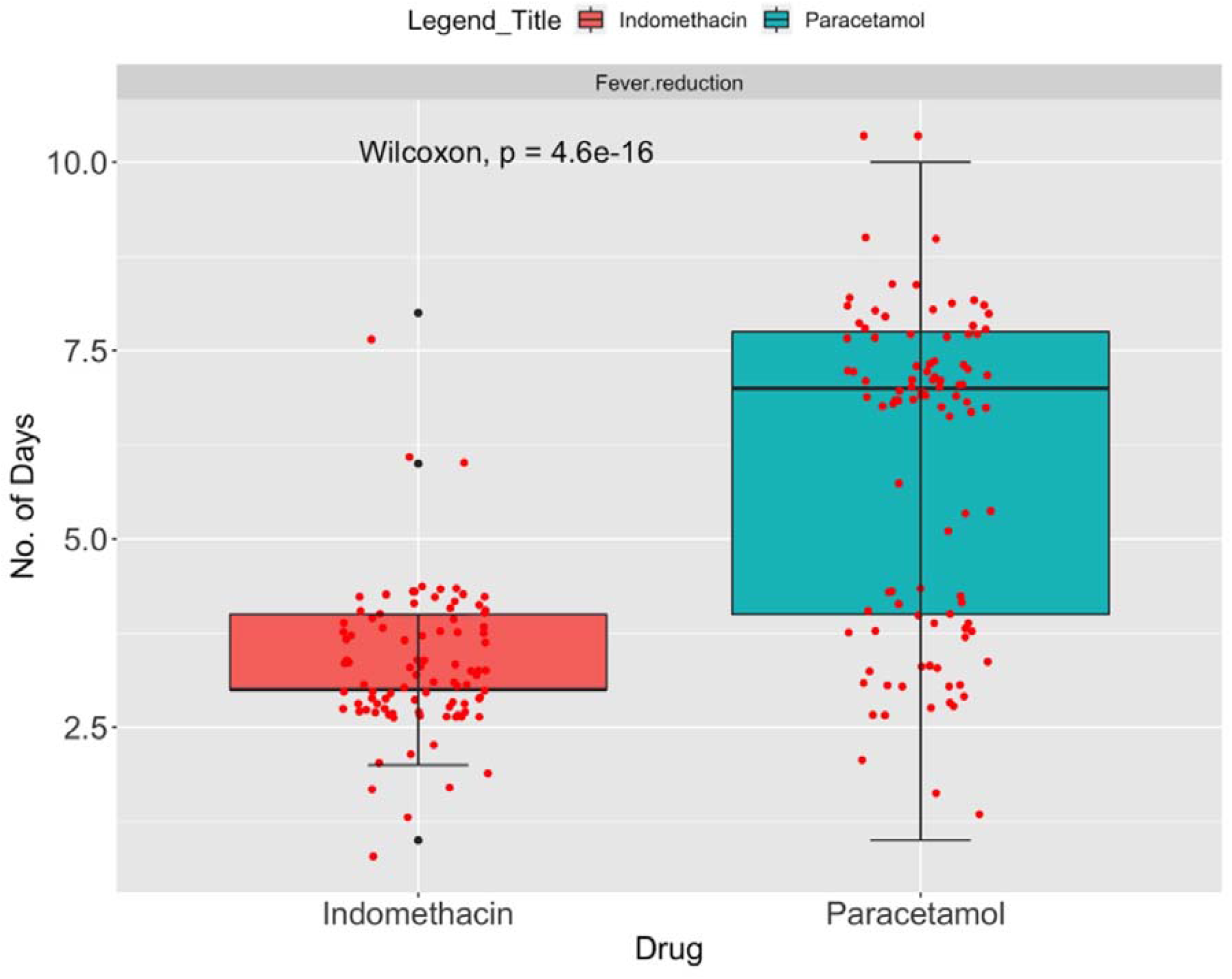
Number of days for Afebrile. N_indomethacin_ = 95; N_Paracetamol_ = 98

**Fig. 3b.**
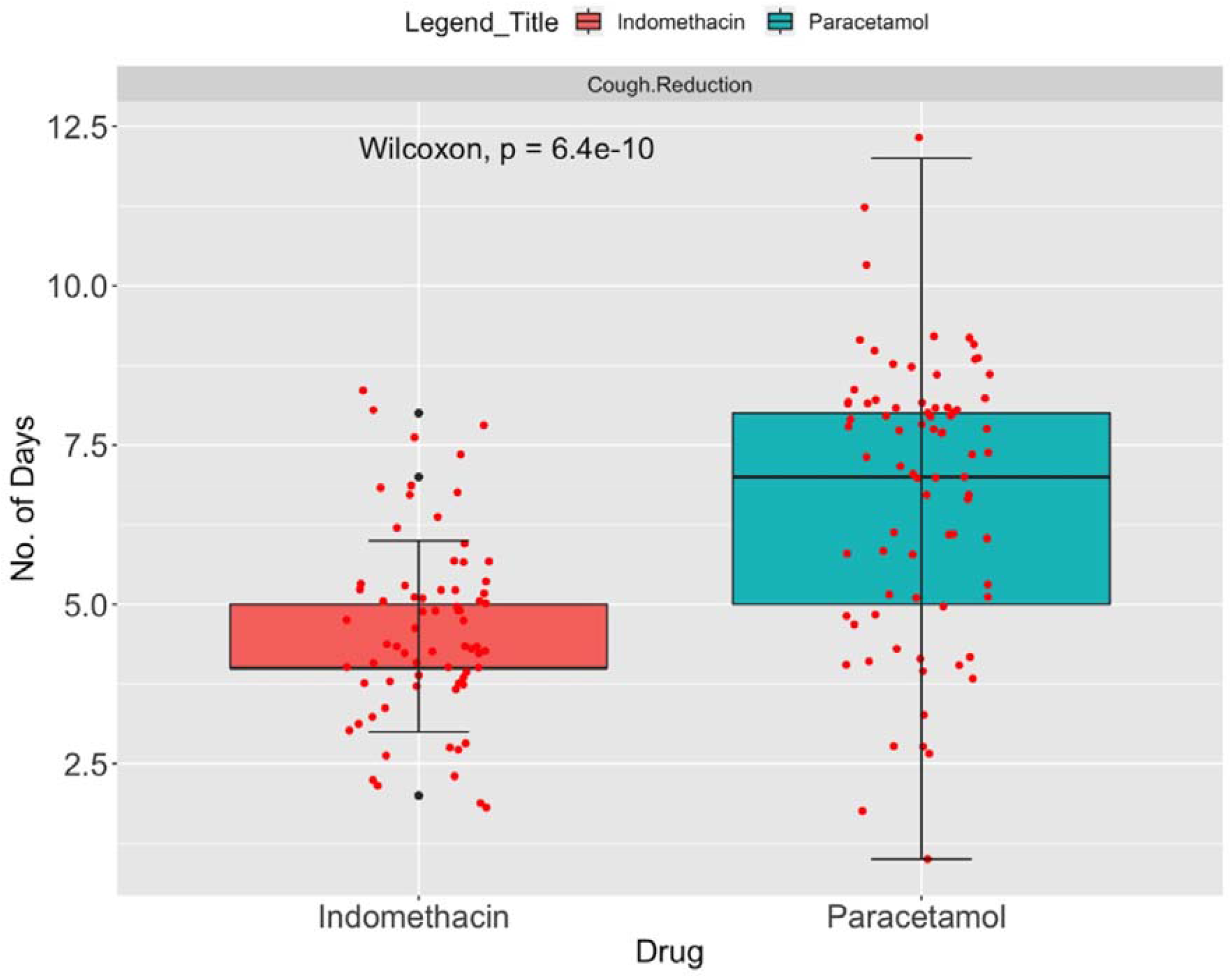
Number of days of days for cough reduction. N_indomethacin_ = 70; N_Paracetamol_ = 75

**Fig. 3c.**
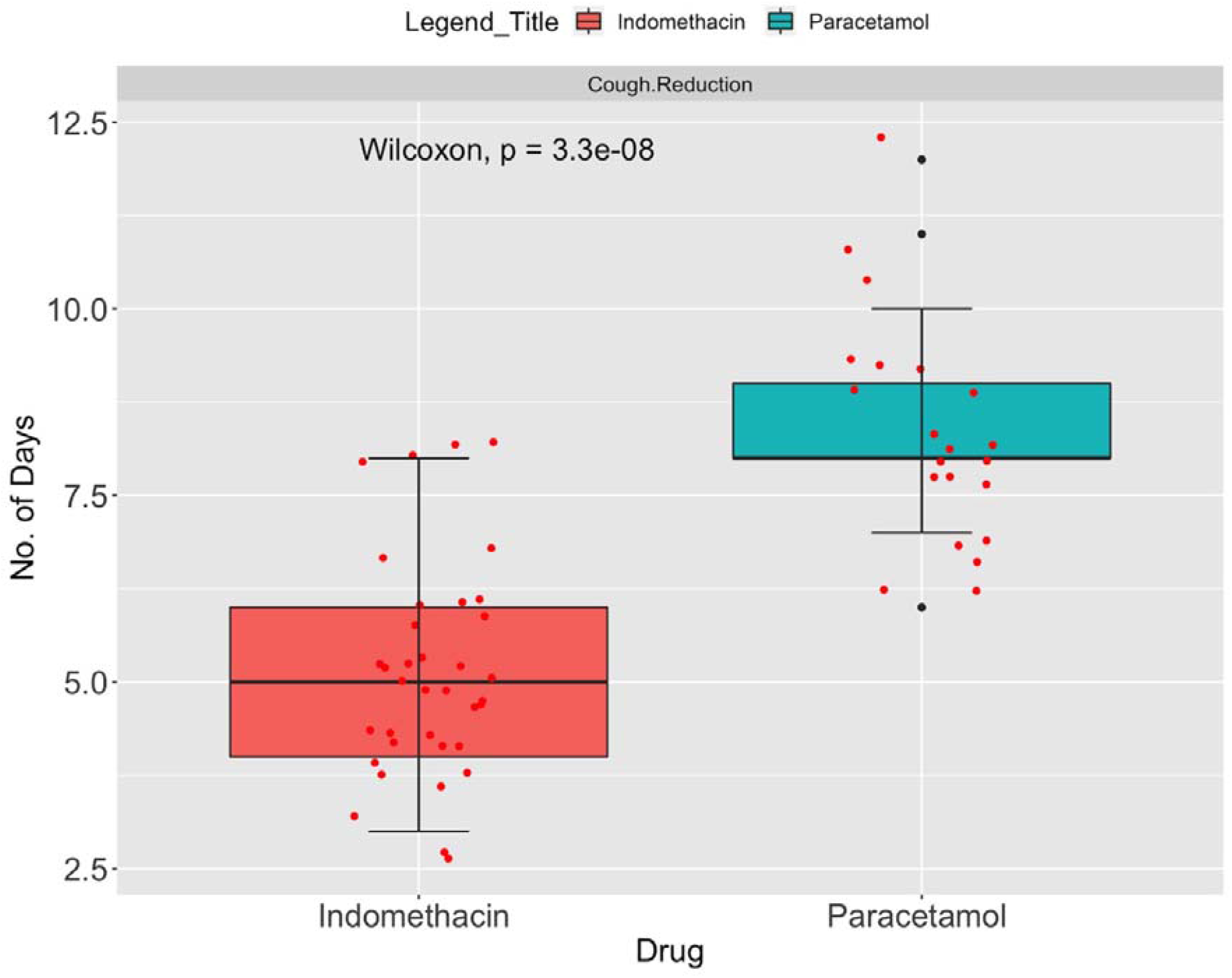
Number of days of days for cough reduction – Cough on admission 7 to 10

**Fig. 3d.**
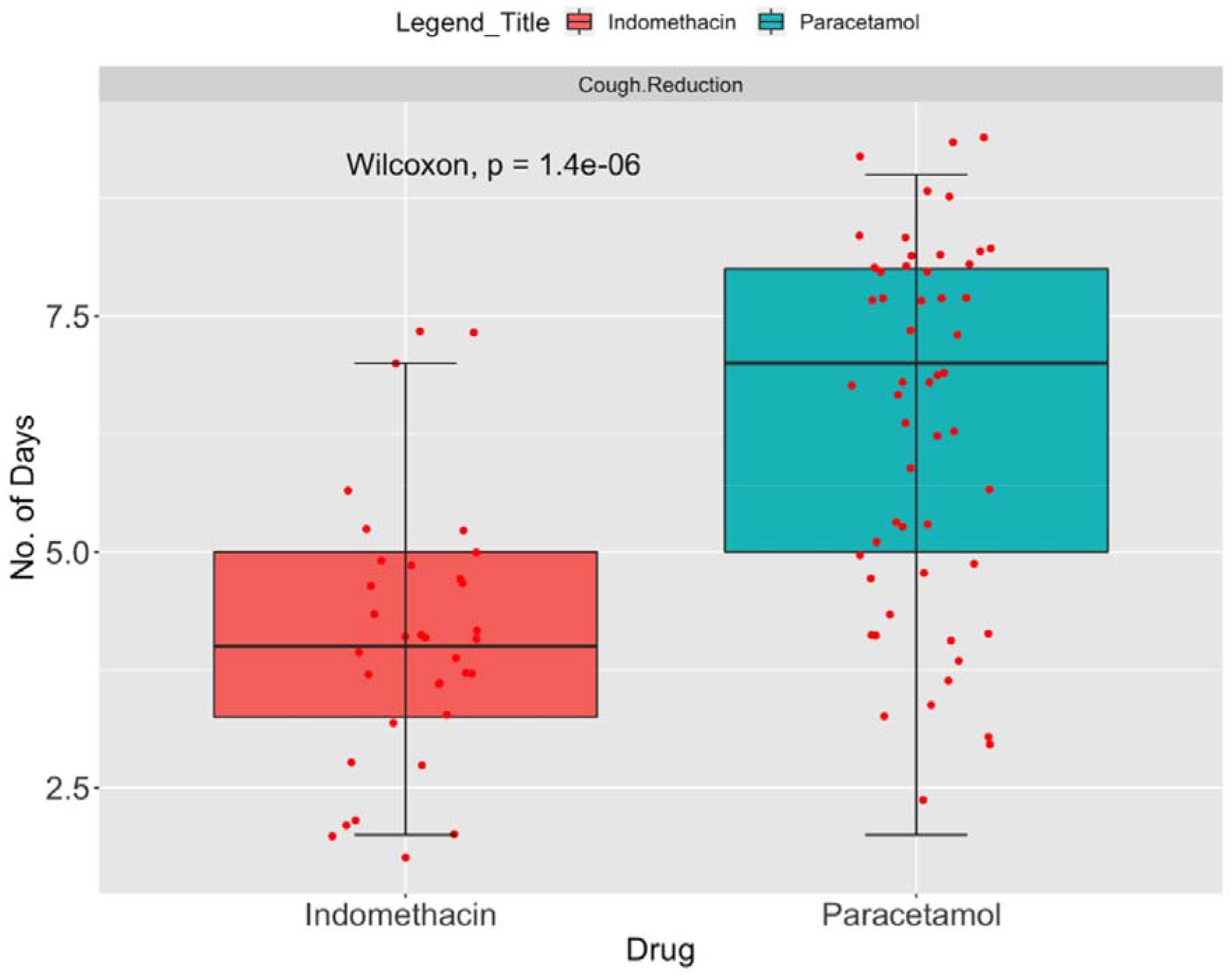
Number of days of days for cough reduction – Cough on admission 2 to 6

**Fig. 3e.**
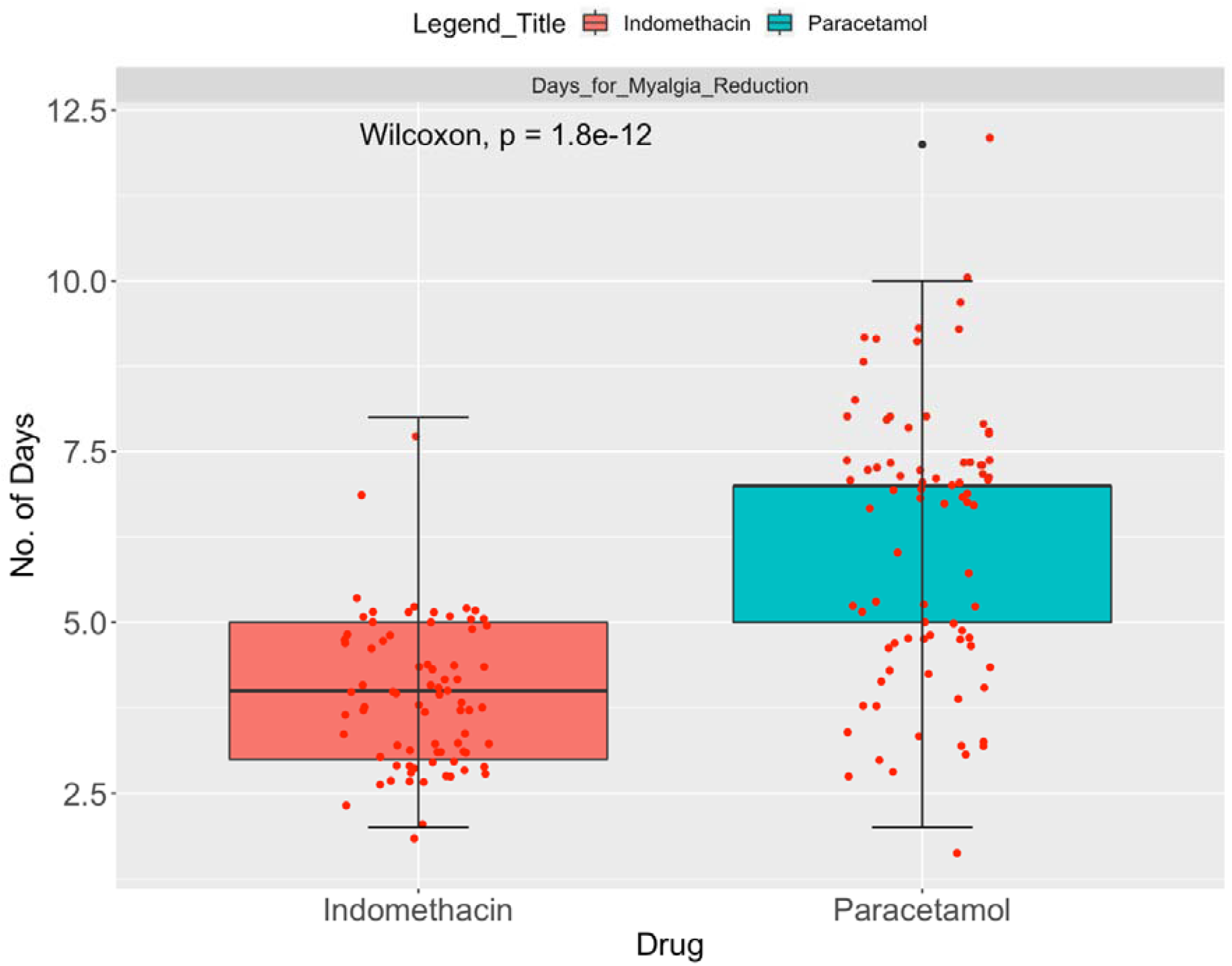
Number of days of days for myalgia resolution. N_indomethacin_ = 77; N_Paracetamol_ = 82

**Table 3.**
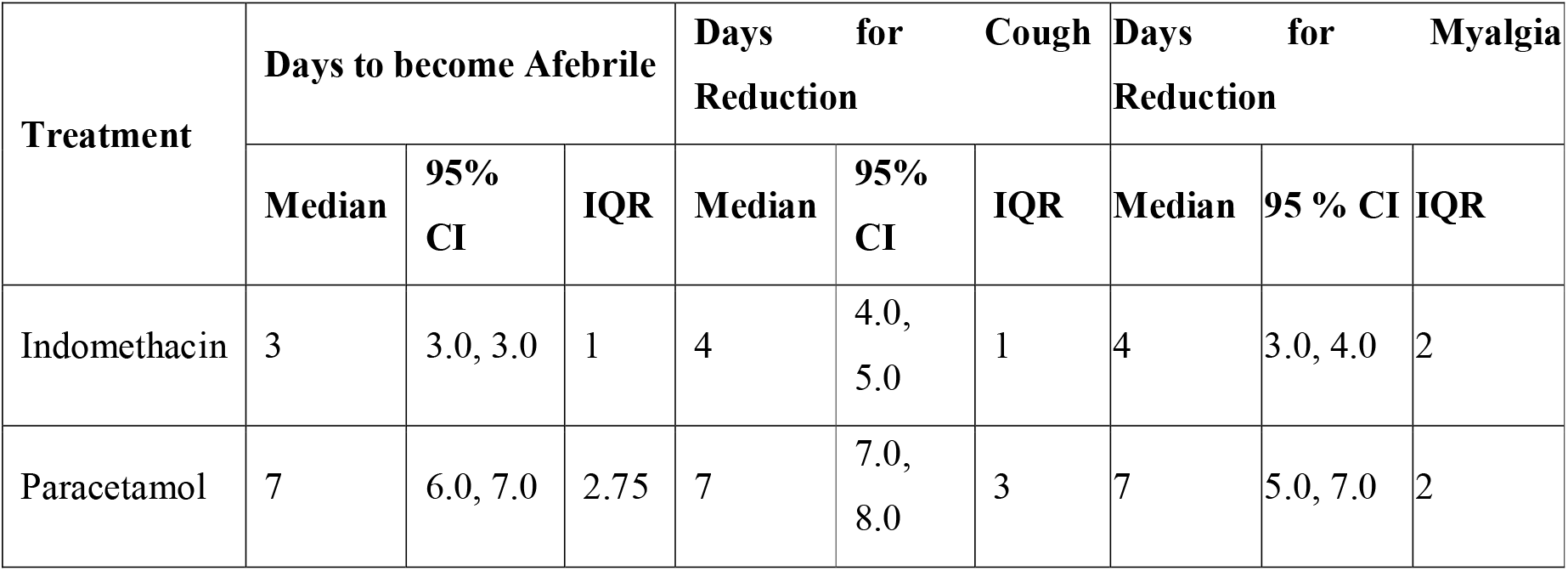
Symptomatic relief due to various treatments

| Treatment | Days to become Afebrile |  |  | Days for Cough Reduction |  |  | Days for Myalgia Reduction |  |  |
| --- | --- | --- | --- | --- | --- | --- | --- | --- | --- |
|  | Median | 95% CI | IQR | Median | 95% CI | IQR | Median | 95 % CI | IQR |
| Indomethacin | 3 | 3.0, 3.0 | 1 | 4 | 4.0, 5.0 | 1 | 4 | 3.0, 4.0 | 2 |
| Paracetamol | 7 | 6.0, 7.0 | 2.75 | 7 | 7.0, 8.0 | 3 | 7 | 5.0, 7.0 | 2 |

Further results are provided in the supplementary appendix. They provide the Kaplan–Meir estimator for the time for relief of three symptoms, namely fever, cough and myalgia in Fig. S1 to S3. Also given in the supplementary appendix are the Cox regression analysis (Tables S1 to S3).

The key question is the number of patients desaturating (SpO_2_ ≤ 93) in both arms. The patients were admitted with a median SpO_2_ of 95 (IQR = 1) and SpO_2_ of 96 (IQR = 1) in the indomethacin arm and paracetamol arm, respectively, with a minimum SpO_2_ of 94. Out of 107 patients, 20 desaturated in the paracetamol arm, while none desaturated in the indomethacin arm. No patient required high-flow oxygen; prone position and occasional low-flow oxygen was sufficient. In the indomethacin arm, saturation improved in one or two doses. The results are presented in Fig. 4.

**Fig. 4.**
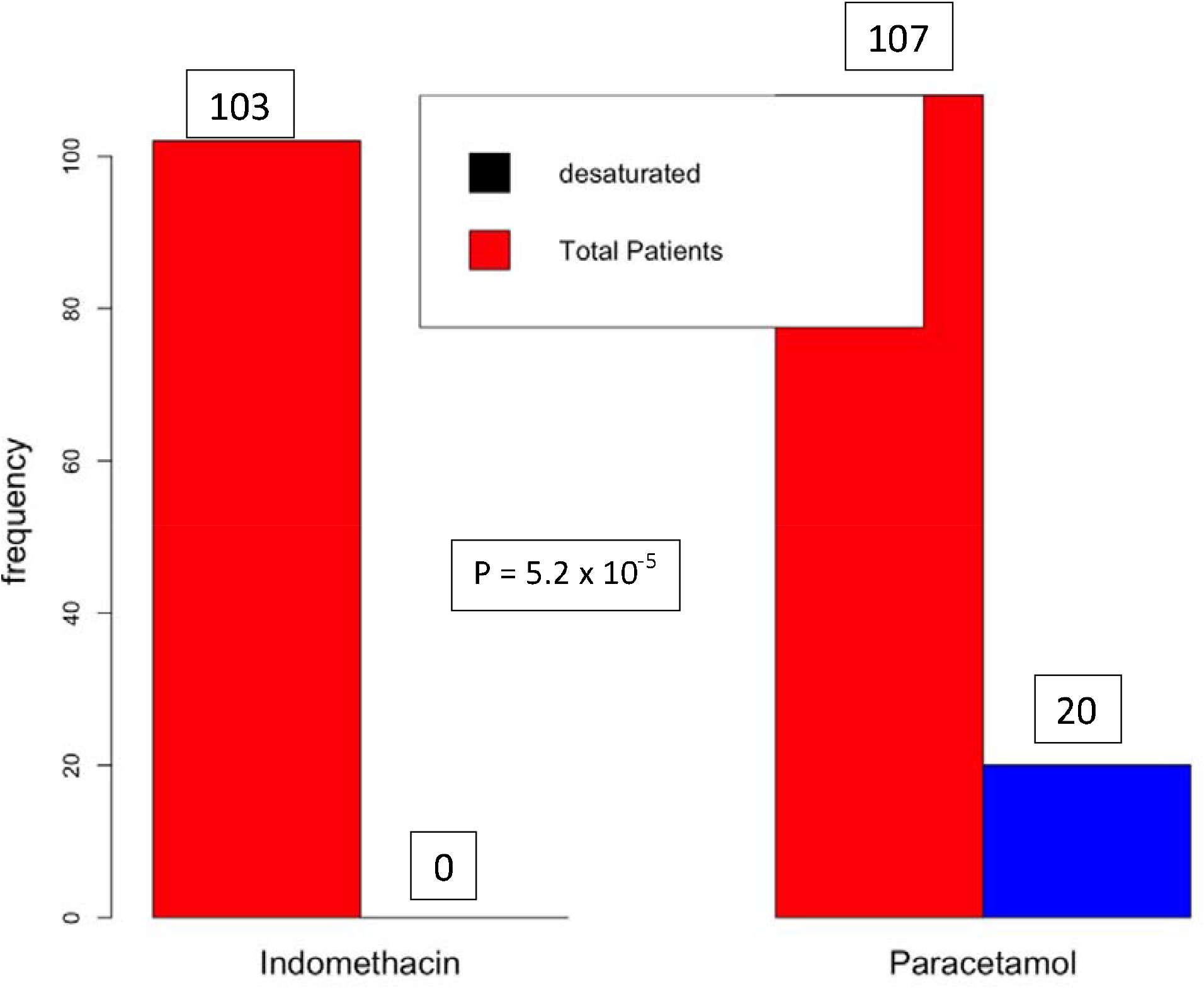
Number of Patients Desaturated

CRP is a well-known inflammatory marker, implicating covid-19 as a severe disease. According to Liu et al.,[16] a CRP score greater than 41.8 may lead to a severe disease. Fig. 5 shows the relationship between the CRP level on admission and the decrease in the CRP level on the seventh day. Fig. 6 shows the relationship between CRP on admission with a value greater than 41 and a reduction in CRP. Although the information in Fig.6 is available in Fig.5, the separation provides clarity of data.

**Fig. 5.**
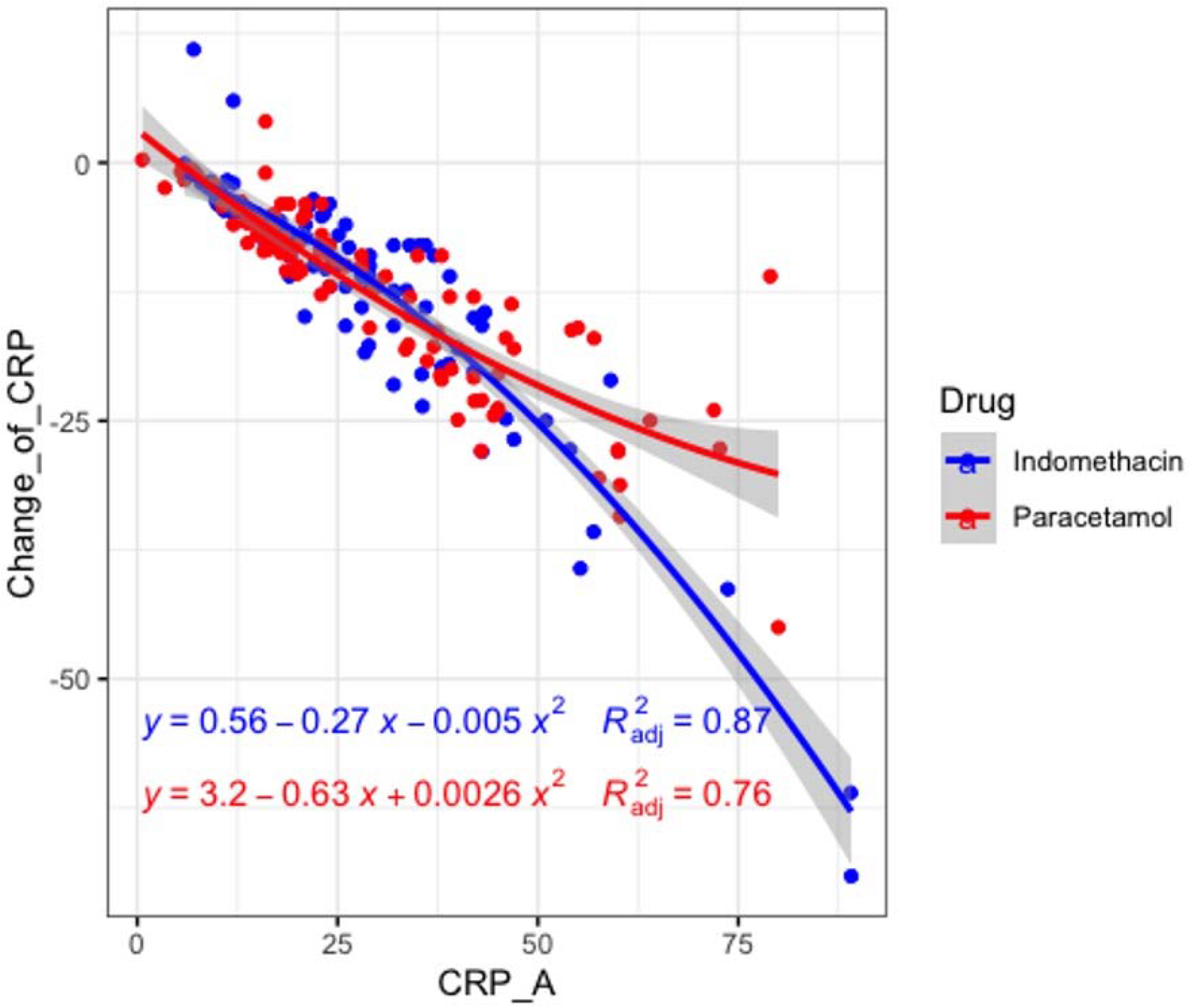
Change in CRP vs. CRP on admission

**Fig. 6.**
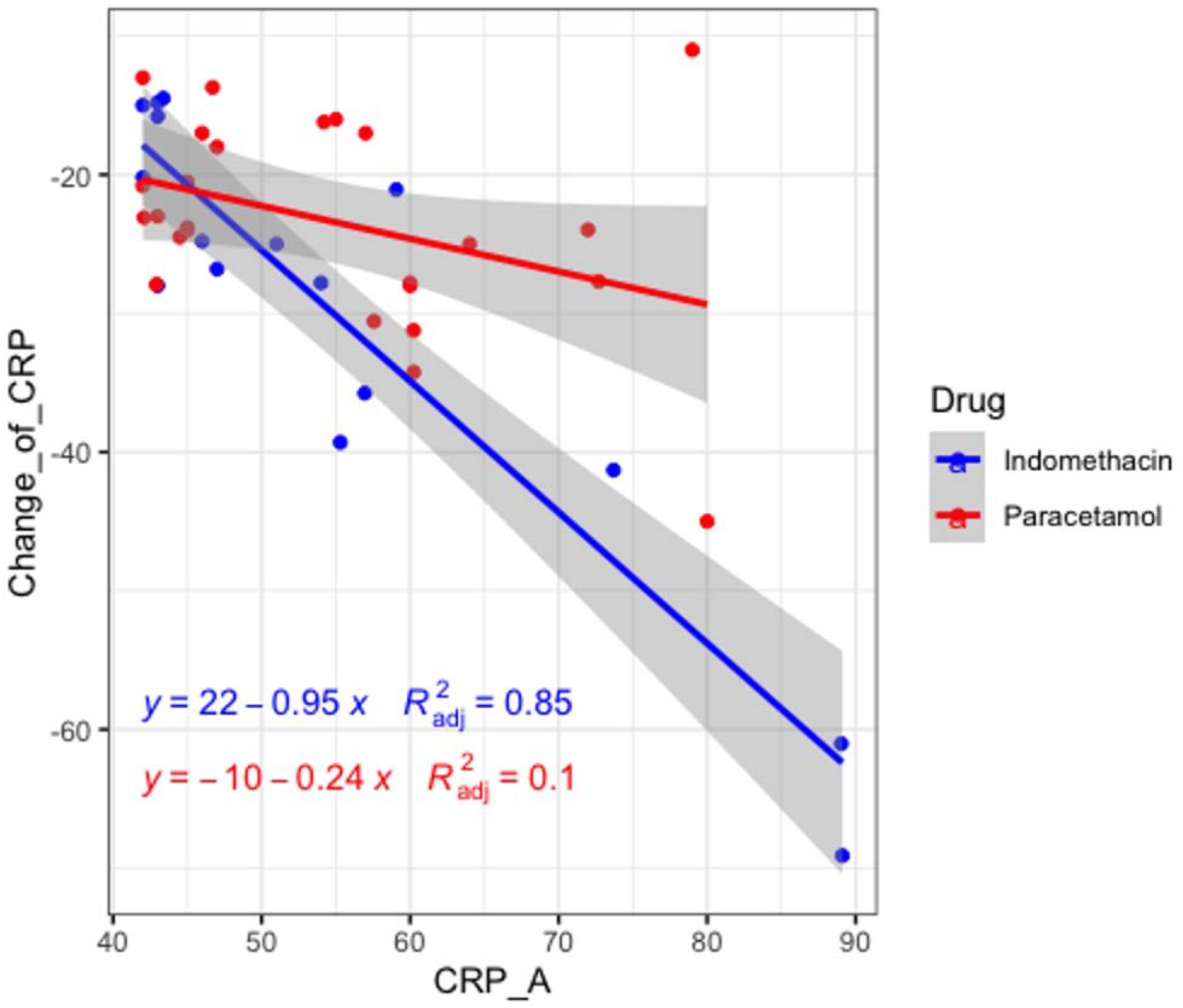
Change in CRP vs. CRP on admission – A closer look of higher CRP on admission

The results of the RT-PCR test conducted on the seventh day are presented in Fig. 7. As can be seen from the figure, more patients became RT-PCR negative on the seventh day in the indomethacin arm although the p-value was 0.42.

**Fig. 7.**
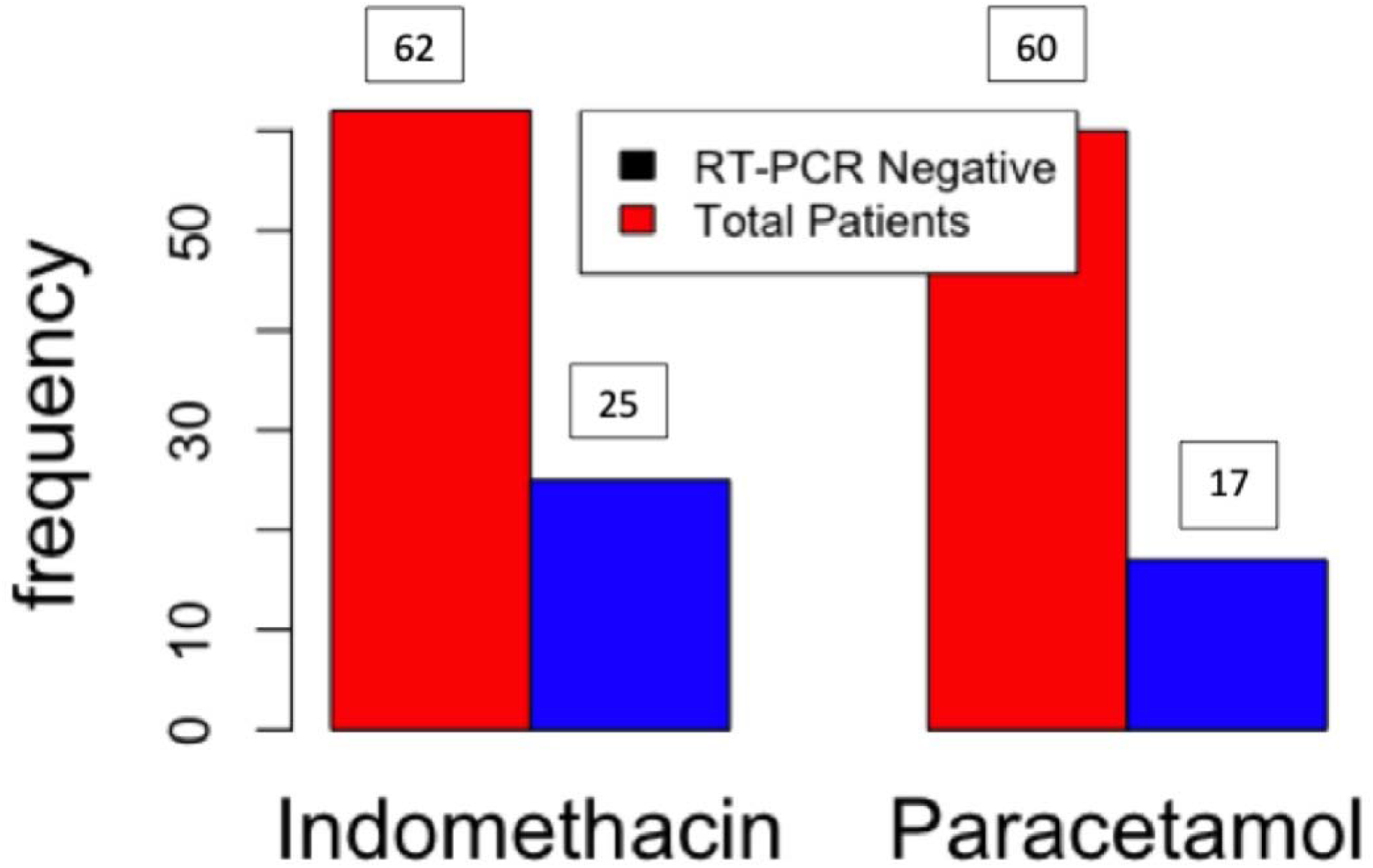
RT-PCR at the seventh day in both the arms

Post-hoc calculations based on actual results yielded a marginal power of 0.99.

### Safety Analysis

Indomethacin was approved in 1965. However, there have been concerns regarding its safety [27]. The number of prescriptions for indomethacin were 2.16 million [28] in 2018, in the US alone. Patients were tested for serum urea and creatinine, SGOT and SGPT before and after the treatment, and the results are shown in Fig. 8 and Fig. 9.

**Fig. 8.**
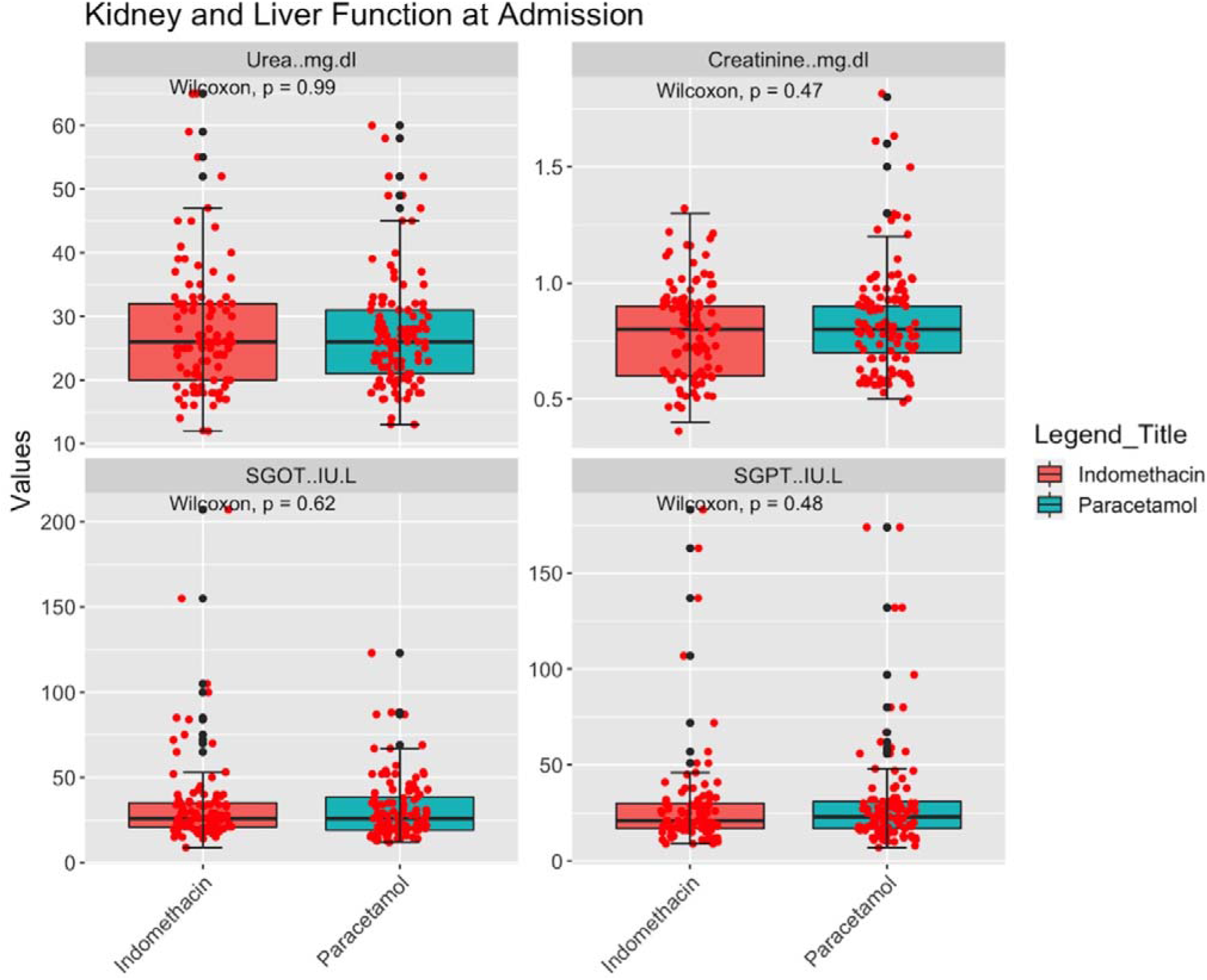
Kidney and Liver function test on admission

**Fig. 9.**
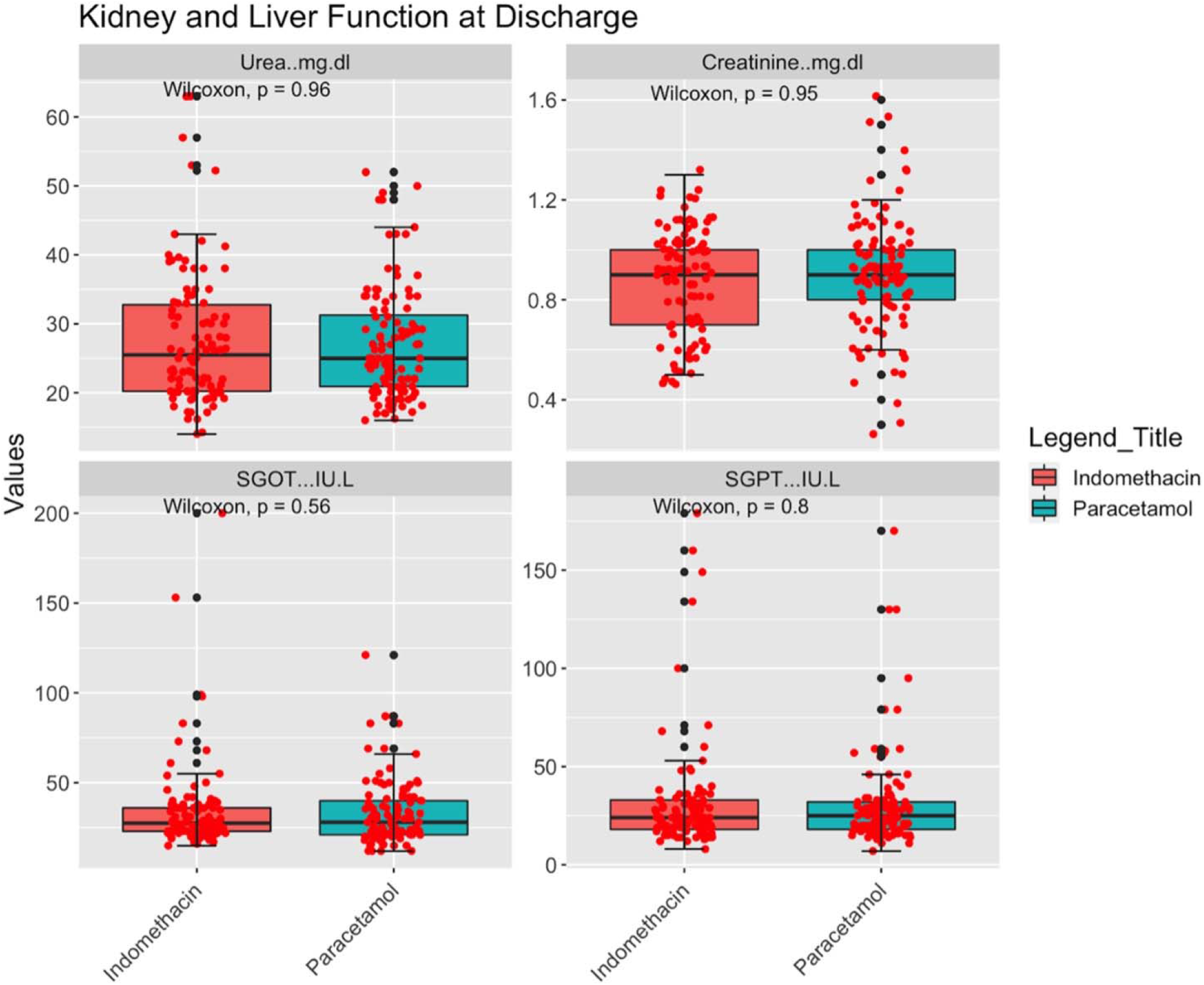
Kidney and Liver function test on discharge

Fig. 8 and Fig. 9 show statistical similarity between both arms. Neither the liver nor kidney function deteriorated after treatment in either arm. Neither the Patients nor the attending physicians did not report any other side-effects.

## Discussion

The primary aim of the study was to understand the efficacy of indomethacin in preventing desaturation (SpO_2_ ≤ 93) and deterioration in mild and moderate covid-19 patients and compare this with a paracetamol-based arm. The secondary aim was to evaluate symptomatic relief in indomethacin patients compared to paracetamol patients. The results are striking: Indomethacin arm patients did not develop desaturation, while nearly 20 per cent of the paracetamol arm patients developed desaturation. When the SpO_2_ level dips below 93, we managed the patient with either low-flow oxygen or by placing them in prone position to enhance breathing. No patient in this study showed further deterioration. Notably, SpO2 improved just after one or two doses in the indomethacin arm. Patients with a marginal SpO2 level of 94 showed an improvement. At the end of the seventh day, 13 patients in the paracetamol arm were at a SpO2 level of 94. In the indomethacin arm, only two patients had SpO2 levels of 94, while 97 of the 103 patients had a higher SpO2 level, higher than 97. In the paracetamol arm only 51 patients (out of 107 patients) had a SpO_2_ level higher than 97.

Symptomatic relief was even more salient. The median time for becoming afebrile was three and seven days in the indomethacin and paracetamol arm, respectively. The median time for cough reduction was four days and seven days in the indomethacin and paracetamol arm, respectively. 59 out of 107 patients in the paracetamol arm had fever on the seventh day while none of the indomethacin arm patients did. 49 of 75 patients taking paracetamol took seven or more days to recover from cough; only nine out of 70 patients in the indomethacin arm took seven days or more to recover and they were at level 2 in the ordinal scale. No patient in the indomethacin arm required continuation of indomethacin after the five-day regimen. On the other hand, patients who were not afebrile after the treatment regimen of five days in the paracetamol arm were continued with the paracetamol at the discretion of the treating physician. One of the most important conclusions came from analysing the IQR. Fig. 3a and 3b show a very thin IQR band for fever and cough reduction in indomethacin patients, along with a small error bar, compared to paracetamol patients. A marginally broader IQR brand in myalgia may indicate the subjective nature of relief.

Kaplan-Meir estimator reinforces the conclusion of the benefit of indomethacin strikingly. All the figures in the supplementary appendix show a big gap between the two treatments. Cox regression results indicate that, the hazard ratio with the treatment of indomethacin is highly significant compared to other covariates. Indomethacin improves the recovery time by 85 to 90% for symptomatic relief. The other covariates that have an effect on cough recovery are the CT Score on admission and cough on admission. Fever reduction and myalgia resolution are not affected by the covariates.

The results are similar to our earlier study which used propensity score matching [21]. In that study, the median time for becoming afebrile, cough reduction, and myalgia relief in the indomethacin arm was four, three, and four days, respectively. However, the median in the paracetamol arm was seven and eight and 6.5 days, respectively.

We monitored CRP on admission and discharge. Indomethacin is very effective in reducing CRP in patients with higher CRP levels on admission (> 41 mg/L). The R2 value for indomethacin (0.85) was much higher compared to paracetamol (0.1). Thus, we can conclude that the consistency of indomethacin in reducing inflammation is very high.

A fourteen-day follow-up further revealed the efficacy of the drug. In the indomethacin arm, nearly 50% patients had fully recovered compared to 28% in the paracetamol arm. The major complaint of the 50% of patient who took indomethacin was tiredness. Only 14% of the patients had myalgia while 10% complained of joint pain. On the other hand, in the paracetamol arm, the recovery was slower with 47% complaining of myalgia, 48% of tiredness, 39% of joint pain and 33% complaining of other ailments.

Alkotaji et.al. [29] had hypothesised the importance of indomethacin in reducing cough and myalgia through its inhibitive action on bradykinin and the mechanism and ill-effects of bradykinin are well documented. The theory proposed in this study explains the difference in symptomatic relief between the two groups.

We had hypothesised that early symptomatic relief is important for recovery and prevents desaturation. 17 of the 20 patients who desaturated had fever beyond seven days and eleven had cough beyond seven days. The fourteenth-day state of the patients also reflected this.

The viral load reduction was better for indomethacin. However, the difference was not significant. Seven days may be too early for an RT-PCR test.

## Limitation of the Study

The limitation of the study is that indomethacin was administered with standard care and not as a stand-alone treatment. With this result, indomethacin may be tried as a stand-alone treatment. This is not a double blinded study. The trial involved only hospitalised patients and not under home isolation.

## Conclusions

The use of indomethacin alongside standard treatment protocol in hospitalised covid-19 patients was associated with marked symptomatic relief and improved oxygen saturation level. We did not observe any adverse effects.

## Data Availability

All data are available as supplementary file

## Supplementary Data

Anonymised Patient data are given in supplementary data file 1. If further data is required please contact the corresponding author.

## Acknowledgement

The authors acknowledge the generous funding for this study by Mr. Kris Gopalakrishnan, Alumnus, Indian Institute of Technology Madras.

## Supplementary Appendix

**Fig. S1.**
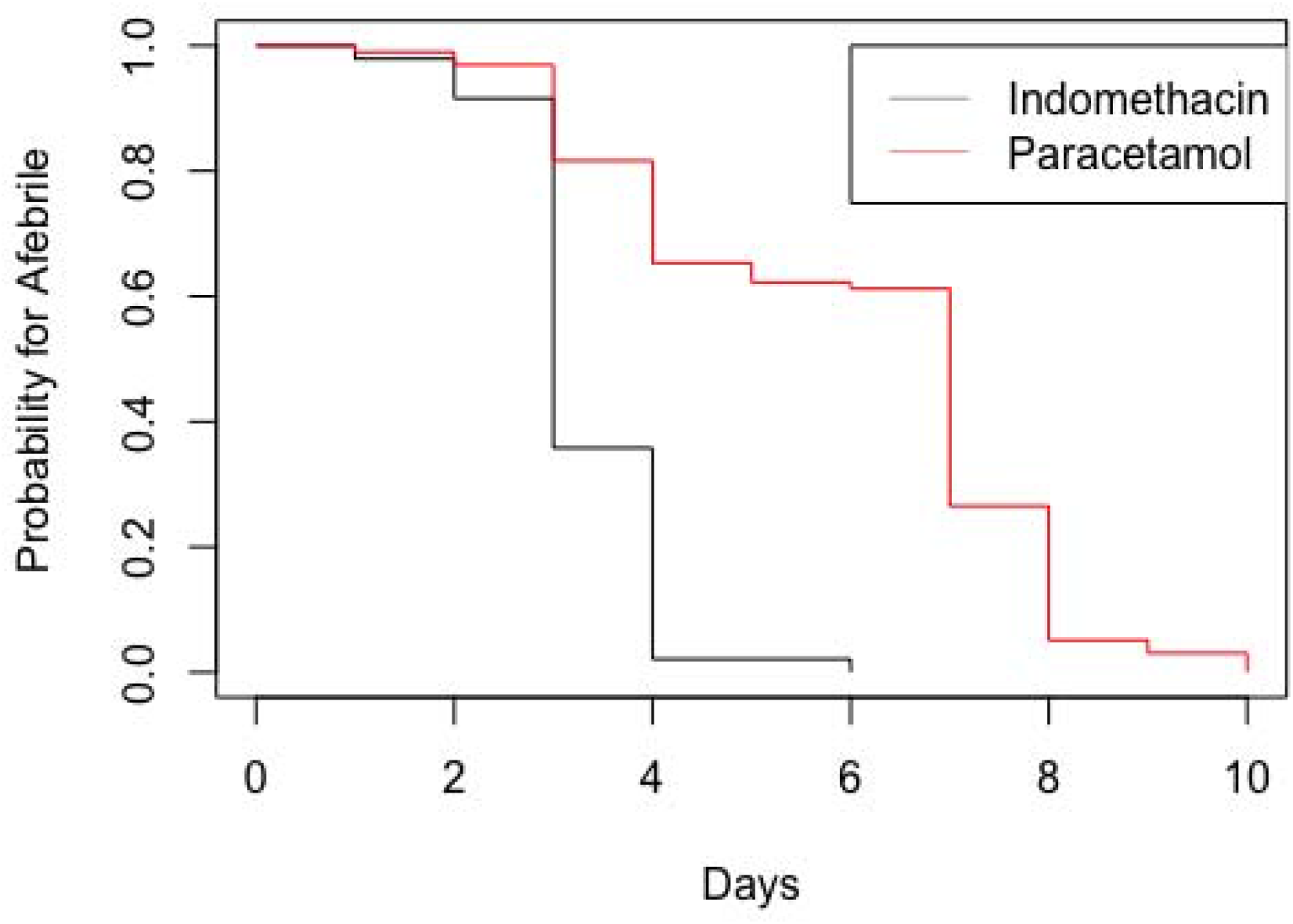
Kaplan-Meir estimate for fever reduction

**Table S1:** Cox Regression results for Fever with either indomethacin or paracetamol treatment; N=193; N_indomethacin_ = 95; N_Paracetamol_ = 98

| Covariates | Reg. Coeff | exp(coef) | z | Pr(>\|z\|) | Lower 0.95 | Upper 0.95 |
| --- | --- | --- | --- | --- | --- | --- |
| Treatment | -2.56955 | 0.07657 | -8.928 | <2e-16 | 0.04356 | 0.1346 |
| Age | 0.013903 | 1.014 | 1.463 | 0.1436 | 0.99528 | 1.0331 |
| CT Score | -0.090987 | 0.91303 | -1.724 | 0.0848 | 0.82328 | 1.0126 |
| Gender | -0.195889 | 0.822104 | -0.878 | 0.3799 | 0.53093 | 1.273 |
| CRP on admission | 0.005697 | 1.005713 | 0.876 | 0.3812 | 0.99297 | 1.0186 |
| Comorbidity | -0.229415 | 0.794999 | -1.21 | 0.2262 | 0.54827 | 1.1528 |

**Fig. S2.**
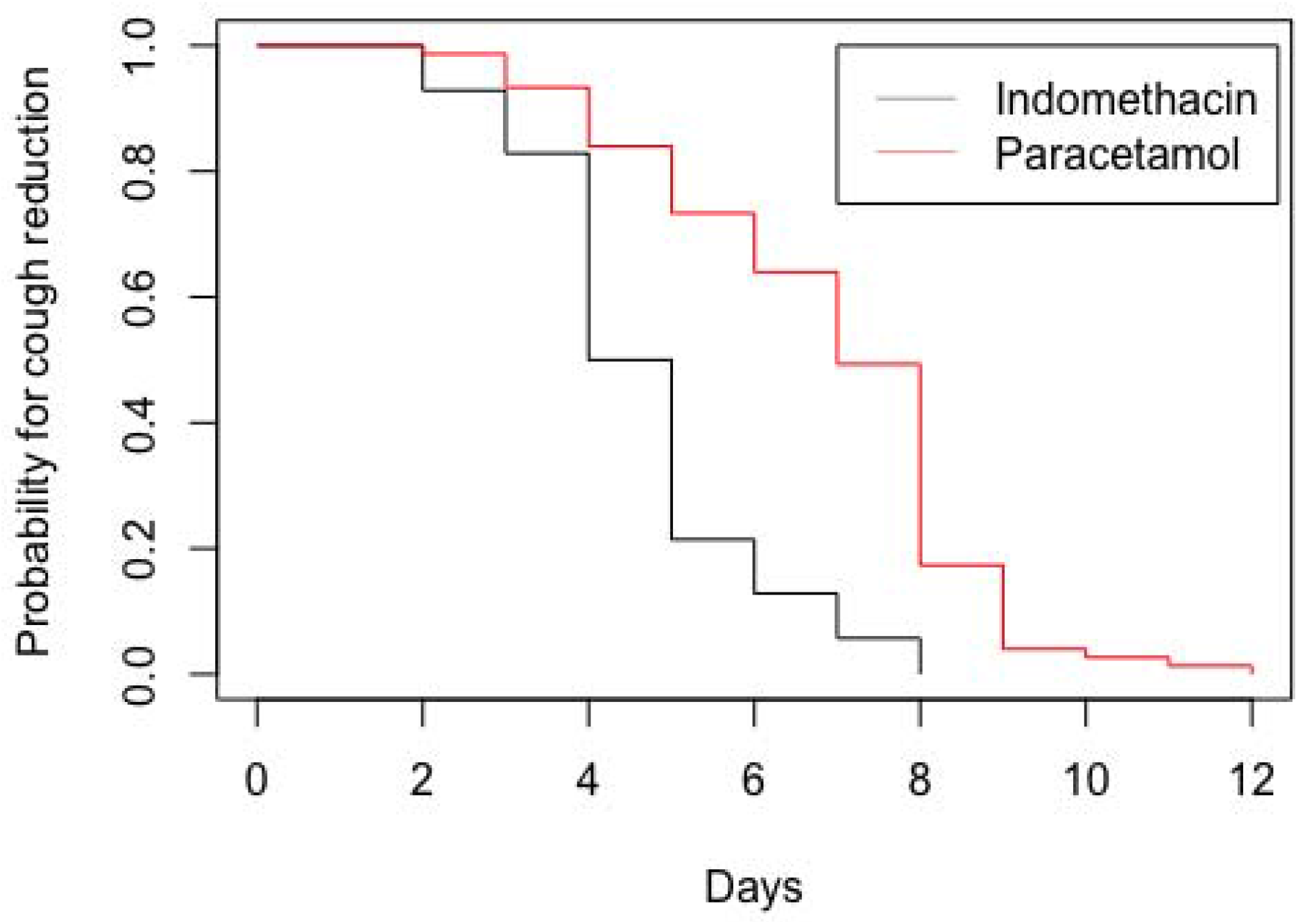
Kaplan-Meir estimate for cough reduction

**Table S2:** Cox Regression results for Cough with either indomethacin or paracetamol treatment; N=145; N_indomethacin_ = 70; N_Paracetamol_ = 75

| Covariates | Reg. Coeff | exp(coef) | z | Pr(>\|z\|) | Lower 0.95 | Upper 0.95 |
| --- | --- | --- | --- | --- | --- | --- |
| Treatment | -2.314587 | 0.098807 | -7.82E+00 | 5.48E-15 | 0.0553 | 0.1766 |
| age | 0.012633 | 1.012713 | 1.36 | 0.17381 | 0.9944 | 1.0313 |
| CT Score | -0.164531 | 0.848291 | -2.919 | 0.00352 | 0.7596 | 0.9474 |
| Gender | -0.173516 | 0.840704 | -0.726 | 0.46779 | 0.5263 | 1.3429 |
| CRP on admission | 0.002738 | 1.002742 | 0.392 | 0.69537 | 0.9891 | 1.0166 |
| Comorbidity | -0.030203 | 0.970248 | -1.66E-01 | 0.86838 | 0.6788 | 1.3868 |
| Cough on admission | -0.251273 | 0.77781 | -5.104 | 3.32E-07 | 0.7063 | 0.8566 |

**Fig. S3.**
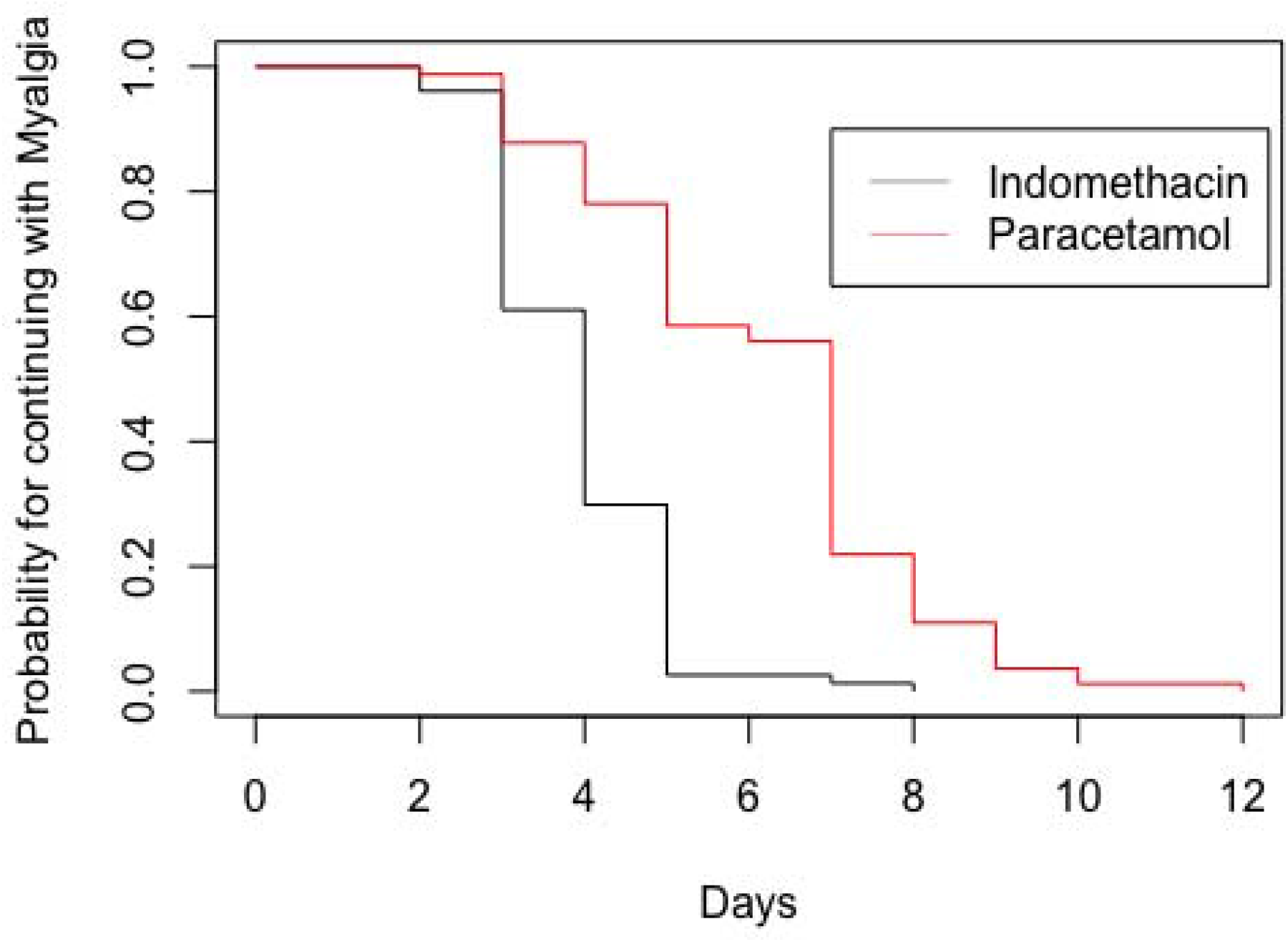
Kaplan – Meir estimate for resolution of Myalgia

**Table S7:**
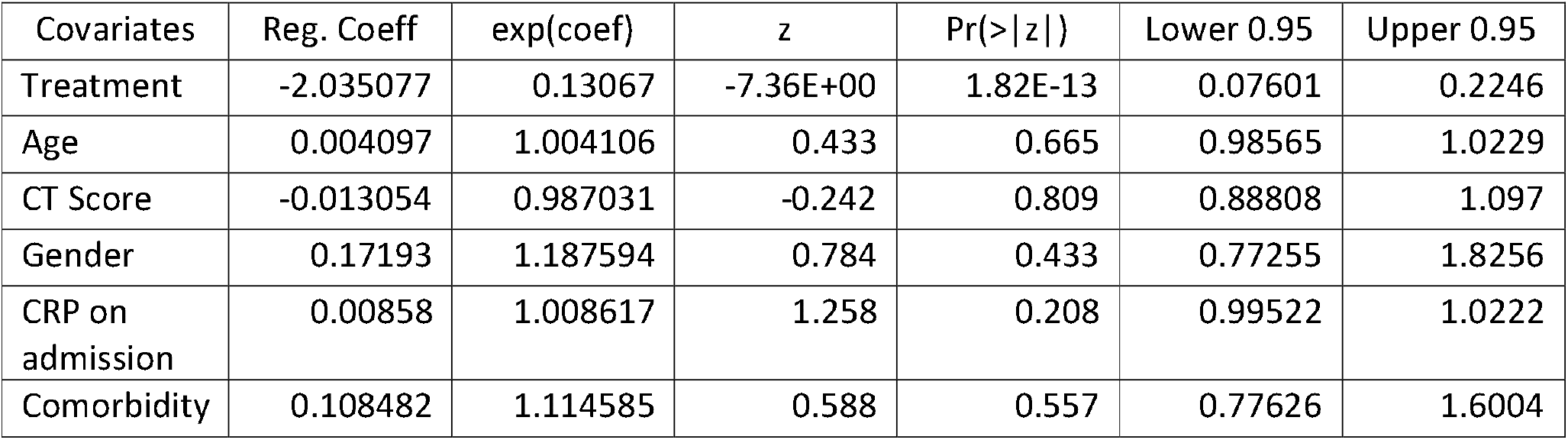
Cox Regression results for Myalgia with either indomethacin or paracetamol treatment; N=159; N_indomethacin_ = 77; N_Paracetamol_ = 82

